# Personalized clinical reference intervals for routine precision medical care

**DOI:** 10.64898/2026.05.28.26354363

**Authors:** Cindy Zhang, Ya-Lin Chen, Attila Jamilov, Elvin Liu, Swati Shree, Barbara D Lam, Brody H Foy

**Affiliations:** Department of Laboratory Medicine & Pathology, University of Washington Medicine, Seattle, USA; Department of Biomedical Informatics & Medical Education, University of Washington Medicine, Seattle, USA; Division of Maternal Fetal Medicine, Department of Obstetrics & Gynecology, University of Washington Medicine, Seattle, USA; Division of Hematology & Oncology, Department of Medicine, University of Washington Medicine, Seattle, USA; Clinical Research Division, Fred Hutchinson Cancer Center, Seattle, USA; Department of Bioengineering, University of Washington, Seattle, USA; Brotman Baty Institute for Precision Medicine, Seattle, USA

## Abstract

Most routine clinical markers are interpreted using population-based reference intervals, despite being regulated around patient-specific homeostatic setpoints. This mismatch obscures physiologic shifts, inhibiting detection of early disease signatures. Here, we develop a novel Bayesian inference method that adaptively constructs personalized reference intervals using each patient’s existing health records. In analysis of >100 million lab tests in >800,000 patients, these personalized intervals can be accurately constructed with only minimal prior data, meaning this method can be applied near universally. We show that across 43 common lab markers, patient setpoints are strongly associated with future morbidity, with signal strength increasing as more test data is collected. Deviation from personalized reference intervals provides strong and novel risk signatures across diverse disease states, including hypothyroidism, hematologic cancers, kidney disease, and pregnancy complications. Importantly, personalized reference intervals capture a different risk signature to existing population-based approaches, with the highest risk patients being those who deviate from both intervals simultaneously. In a targeted clinical use case study of iron infusion, use of personalized reference intervals greatly improved prediction of treatment efficacy and allowed precise tracking of treatment responses. Our results illustrate how existing health records can be used to construct personalized benchmarks for nearly all common clinical tests, driving a new paradigm for precision laboratory medicine.

## Introduction

Clinical laboratory testing is one of the frontlines of medicine, often providing the first quantitative assessment of a presenting patient. Routine tests such as the blood count and metabolic panel are used across nearly all areas of medicine^1,2^ to provide rapid insights into hematologic, immunologic, metabolic, renal, and cardiovascular function. Most of the resulting panel markers are used as general monitors of physiologic state, rather than as diagnostics for a singular disease. As such, results are typically interpreted using population reference intervals (popRIs), traditionally set as the central 95% of values from a healthy reference population^3^. These intervals provide a standardized and scalable framework for clinical screening across medical fields.

While easy to implement, popRIs are often unreflective of individual physiology and health history^4,5^. Many common markers, such as blood cell traits (hemoglobin, white cell count, etc.) and metabolic markers (creatinine, sodium, etc.) are highly heritable and polygenically regulated^6^, meaning what is “normal” will vary based on patient genetics. Similarly, marker regulation may shift longitudinally due to lifestyle factors (weight, smoking, etc.^7,8^), physiologic events (e.g., menopause^9^, pregnancy^10^, etc.), development of chronic comorbidities (e.g., chronic kidney disease^11^, polycythemia vera^12^, etc.), or clinical interventions (medication^13^, surgery, etc.). Consequently, the use of popRIs can inappropriately conflate population- and patient-level abnormality for a given marker. On one side, clinically relevant deviations for a patient may remain within the popRI, delaying disease detection. On the other side, stable biologic fluctuations for a patient whose marker setpoint is near a reference limit may be continually flagged as abnormal, leading to unnecessary clinical workups^14^. Despite longstanding recognition of these limitations^15^, popRIs remain the dominant operational reference framework in nearly all medical fields due to the lack of a clear alternative.

With the modern rise of electronic health records (EHRs), many patients now have extensive prior testing histories available, particularly for routine markers. In recent work, we showed how longitudinal EHR data can be used to estimate setpoints for the complete blood count (CBC). While novel, the Gaussian mixture model (GMM) used in that study relied on extensive prior testing histories, making it clinically impractical for most patients. To leverage personalized regulation clinically, there is a need for an adaptive framework that can refine estimates of patient homeostasis as their health record history expands.

To address this, we present a Bayesian inference method for constructing patient-specific, personalized reference intervals (perRIs). We show that perRIs can accurately predict future patient test values for 43 of the most common clinical blood markers. Associated setpoint estimates are strongly associated with diverse morbidities and enhance interpretation of new test results. perRIs provide complementary information to popRIs, with joint use of both approaches greatly enhancing disease detection and treatment response prediction. Our approach provides a simple and adaptive framework for routine precision laboratory medicine.

## Results

We studied a multi-decade, longitudinal database of health records from a large, multi-site medical center, spanning 814,371 patients, 111,498,837 laboratory tests and 43 markers over 17 years. Definitions of each marker and cohort summary statistics are given in **Table 1**. Marker measurement distributions and individualized setpoint distributions are shown in **Supp. Figs. 1–2**.

**Table 1.** Cohort summary.

| Abbreviation | Marker Name | Units | PopRI | N | Setpoint | Setpoint CV | Age | % Male |
| --- | --- | --- | --- | --- | --- | --- | --- | --- |
| *FER | Ferritin | ng/mL | F: [10, 180]; M: [20, 230] | 30,490 | 125.9 (340.7) | 32.9% (14.0%) | 50.1 (19.0) | 28.4 |
| *GLU | Glucose | mg/dL | [62, 125] | 194,907 | 103.9 (30.8) | 10.7% (3.5%) | 54.3 (16.4) | 46.8 |
| *CRP | High-Sensitivity C-Reactive Protein | mg/L | [0, 10] | 18,289 | 6.0 (8.1) | 535.8% (805.6%) | 53.0 (16.9) | 36.8 |
| *TRIG | Triglycerides | mg/dL | [0, 175] | 134,754 | 138.5 (89.9) | 91.4% (63.1%) | 55.8 (15.0) | 50.4 |
| *TSH | Thyroid-Stimulating Hormone | mIU/L | [0.4, 5] | 70,830 | 2.2 (1.7) | 39.1% (26.6%) | 55.3 (17.1) | 32 |
| A1C | Hemoglobin A1c | % | [4, 5.6] | 40,332 | 6.2 (1.0) | 3.6% (1.2%) | 57.5 (15.0) | 51.8 |
| ALB | Albumin | g/dL | [3.5, 5.2] | 180,791 | 4.3 (0.3) | 4.1% (1.6%) | 54.3 (16.5) | 46.9 |
| ALK | Alkaline Phosphatase | U/L | age-dependent, see Supp. Table 1 | 178,283 | 71.4 (28.7) | 18.0% (8.1%) | 54.3 (16.5) | 46.9 |
| ALT | Alanine Aminotransferase | U/L | F: [7, 33]; M (<49y): [10, 64]; M(>49y): [10-48] | 179,169 | 24.0 (15.1) | 19.5% (9.1%) | 54.3 (16.5) | 46.9 |
| AST | Aspartate Aminotransferase | U/L | [9, 38] | 178,893 | 22.6 (9.5) | 14.3% (5.3%) | 54.3 (16.5) | 46.9 |
| BIL | Total Bilirubin | mg/dL | [0.2, 1.3] | 178,284 | 0.7 (0.3) | 21.3% (9.5%) | 54.3 (16.5) | 46.9 |
| BILD | Direct Bilirubin | mg/dL | [0, 0.3] | 16,329 | 0.2 (0.1) | 22.8% (22.8%) | 56.1 (15.9) | 56.1 |
| BUN | Blood Urea Nitrogen | mg/dL | [8, 21] | 194,629 | 15.6 (5.7) | 11.1% (4.8%) | 54.4 (16.4) | 46.9 |
| CA | Calcium | mg/dL | [8.9, 10.2] | 194,654 | 9.4 (0.3) | 2.3% (0.6%) | 54.4 (16.4) | 46.8 |
| CHOL | Total Cholesterol | mg/dL | [0, 210] | 135,479 | 188.0 (40.5) | 9.5% (4.7%) | 55.7 (15.0) | 50.3 |
| CL | Chloride | mEq/L | [98, 108] | 194,562 | 103.3 (2.4) | 1.5% (0.4%) | 54.4 (16.4) | 46.9 |
| CO2 | Carbon Dioxide | mEq/L | [22, 32] | 194,559 | 27.6 (2.1) | 5.7% (1.7%) | 54.4 (16.4) | 46.9 |
| CRE | Creatinine | mg/dL | F: [0.38, 1.02]; M: [0.51, 1.18] | 193,532 | 0.9 (0.3) | 7.8% (3.1%) | 54.4 (16.4) | 46.8 |
| HB | Hemoglobin | g/dL | F: [11.5, 15.5]; M: [13, 18] | 163,296 | 13.6 (1.4) | 4.1% (1.6%) | 53.2 (17.4) | 41.6 |
| HCT | Hematocrit | % | F: [36, 45]; M: [38, 50] | 164,220 | 40.9 (4.0) | 3.2% (1.2%) | 53.0 (17.5) | 41.5 |
| HDL | HDL Cholesterol | mg/dL | F: >49; M: >39 | 135,438 | 53.0 (15.5) | 8.8% (3.4%) | 55.7 (15.0) | 50.3 |
| IGAP | Anion Gap | mEq/L | [4, 12] | 193,533 | 7.7 (1.6) | 18.4% (6.7%) | 54.5 (16.4) | 46.9 |
| K | Potassium | mEq/L | [3.6, 5.2] | 193,627 | 4.2 (0.3) | 6.0% (1.6%) | 54.4 (16.4) | 46.8 |
| LD | Lactate Dehydrogenase | U/L | [0, 210] | 7,842 | 176.7 (59.9) | 12.6% (5.4%) | 56.2 (18.0) | 57.3 |
| LDL | LDL Cholesterol | mg/dL | [0, 130] | 2,885 | 91.0 (36.6) | 17.2% (10.2%) | 63.6 (13.8) | 56.7 |
| LYMPH | Lymphocytes | x10<sup>3</sup>/uL | [1, 4.8] | 104,253 | 1.9 (0.9) | 25.5% (11.6%) | 55.2 (17.2) | 42.6 |
| MCH | Mean Corpuscular Hemoglobin | pg | [27.3, 33.6] | 162,619 | 30.0 (2.3) | 2.1% (1.0%) | 53.2 (17.4) | 41.6 |
| MCHC | Mean Corpuscular Hemoglobin Concentration | g/dL | [32.2, 36.5] | 162,618 | 33.3 (1.1) | 1.4% (0.5%) | 53.2 (17.4) | 41.6 |
| MCV | Mean Corpuscular Volume | fL | [81, 98] | 162,642 | 90.3 (5.7) | 1.8% (0.8%) | 53.2 (17.4) | 41.6 |
| MG | Magnesium | mg/dL | [1.8, 2.4] | 13,356 | 1.9 (0.2) | 4.9% (1.7%) | 58.5 (17.3) | 51.1 |
| MONO | Monocytes | x10<sup>3</sup>/uL | [0, 0.8] | 104,250 | 0.5 (0.2) | 22.7% (9.5%) | 55.2 (17.2) | 42.6 |
| NA | Sodium | mEq/L | [135, 145] | 194,549 | 138.7 (2.0) | 1.1% (0.3%) | 54.4 (16.4) | 46.9 |
| NHDL | Non-HDL Cholesterol | mg/dL | [0, 190] | 134,814 | 134.9 (38.2) | 12.0% (6.6%) | 55.8 (15.0) | 50.4 |
| P | Phosphate | mg/dL | [2.5, 4.5] | 14,119 | 3.5 (0.6) | 9.5% (2.9%) | 57.1 (17.4) | 53.5 |
| PLT | Platelets | $\times 10^3/\mu\text{L}$ | [150, 400] | 162,170 | 244.8 (67.8) | 13.4% (5.8%) | 53.2 (17.4) | 41.6 |
| INR | INR | ratio | [0.8, 1.3] | 16,066 | 1.2 (0.3) | 5.8% (3.2%) | 58.9 (15.1) | 54.9 |
| PT | Prothrombin Time | sec | [10.7, 15.6] | 15,680 | 14.8 (3.0) | 4.9% (1.5%) | 58.7 (15.1) | 55 |
| RBC | Red Blood Cell Count | $\times 10^6/\mu\text{L}$ | F: [3.8, 6]; M: [4.4, 5.6] | 162,547 | 4.6 (0.5) | 4.9% (2.3%) | 53.2 (17.4) | 41.6 |
| RDW | Red Cell Distribution Width Coefficient of Variation | % | [10, 14.5] | 162,391 | 13.3 (1.2) | 4.0% (1.5%) | 53.2 (17.4) | 41.6 |
| NEUT | Total Neutrophils | $\times 10^3/\mu\text{L}$ | [1.8, 7] | 104,251 | 4.1 (1.6) | 21.4% (7.9%) | 55.2 (17.2) | 42.6 |
| TP | Total Protein | g/dL | [6, 8.2] | 177,592 | 7.0 (0.5) | 4.4% (1.3%) | 54.3 (16.4) | 46.9 |
| VITD | Vitamin D (25-OH) | ng/mL | [20.1, 50] | 37,564 | 34.4 (13.0) | 15.0% (7.3%) | 54.3 (17.7) | 32.1 |
| WBC | White Blood Cell Count | $\times 10^3/\mu\text{L}$ | [4.3, 10] | 162,631 | 6.8 (2.0) | 14.2% (4.6%) | 53.2 (17.4) | 41.6 |

*Legend: Setpoint estimates were performed using the first three isolated outpatient tests for each marker. Unless otherwise noted, data are presented as mean(std). \* denotes markers with setpoints fit on a logarithmic scale. PopRI are defined using UWM laboratory reference values.*

### Bayesian inference accurately constructs personalized reference intervals

To estimate perRIs from patient EHRs, we use a Bayesian inference framework. This method starts by assuming each patient’s setpoint and variance are drawn uniformly from the popRI. It then uses Bayes theorem to refine this estimate, using isolated outpatient test results. Representative model fits for platelet count (PLT) are given in **Fig. 1a**, for the Bayesian model, and the prior published GMM model for CBC setpoint estimation^16^. The Bayesian model shows drastically improved predictive fits and calibration to the GMM model and improved variance calibration, particularly in the presence of low data. Averaged across 43 markers, Bayesian perRI estimates had lower prediction error (scaled root mean squared error (RMSE) 1.087 vs 1.171 for GMM, p<0.001, note: theoretical lower limit of scaled RMSE is 1.0) and better calibrated variance (average Kolmogorov-Smirnov [KS] statistic of 0.113 vs 0.237 for GMM, p<0.001) against future outpatient tests (**Fig. 1c**). Model performance remained significantly better even for markers with limited heritability, as measured by the index of individuality (IOI; the ratio of intra- to inter-patient variance for the marker), as well as for markers with lower testing density (**Fig. 1c-d**). Predictive error decreased with increasing individuality while calibration error increased (*ρ* (IOI, RMSE_scaled_ = -0.59; (*ρ* (IOI, KS) = 0.32) (**Fig. 1e**). Bayesian estimates adapted quickly to individual baselines, with low error even after only 1-3 measurements (**Fig. 1f**).

**Figure 1.**
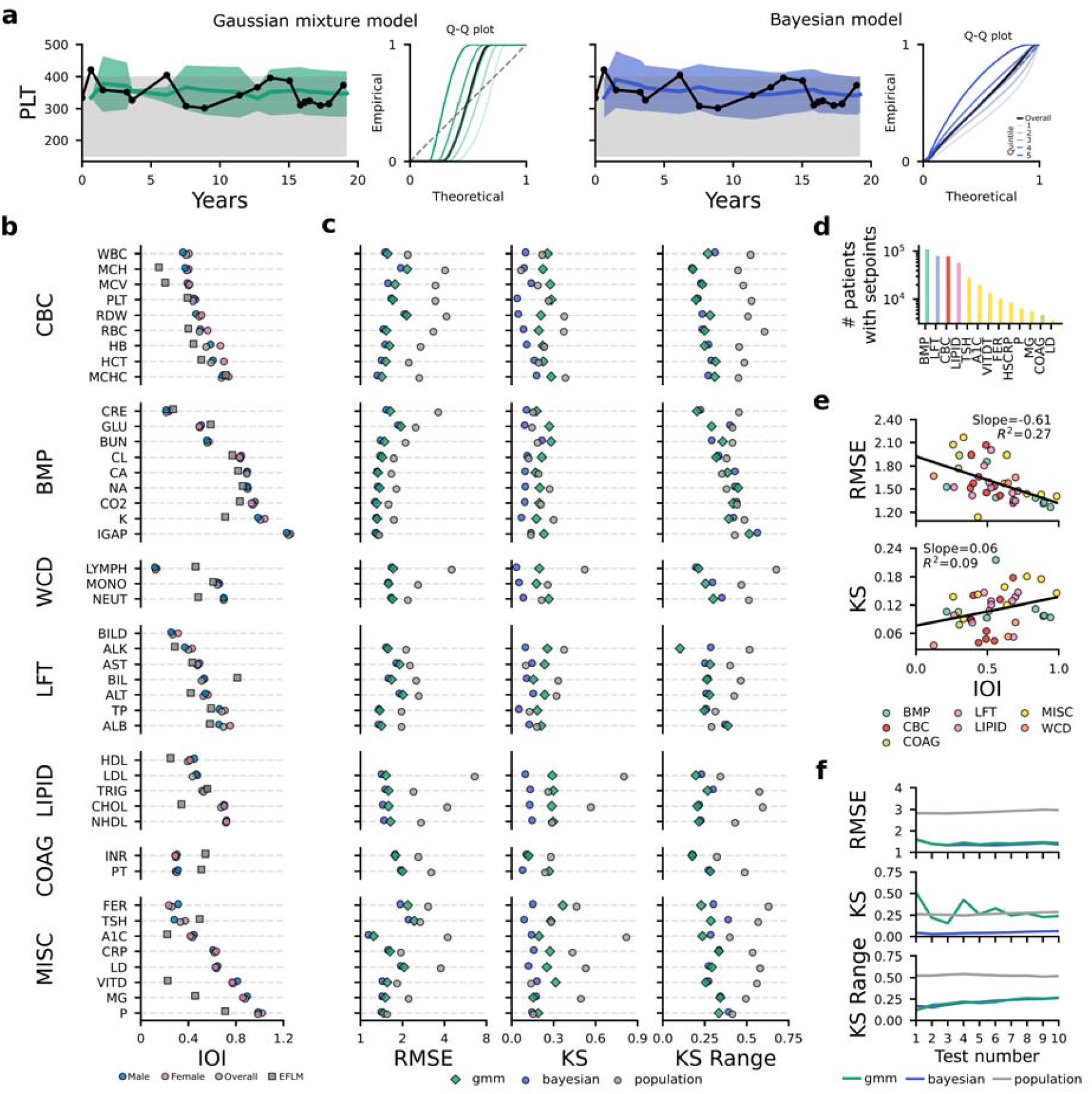
Validation of the Bayesian framework for personalized reference interval estimation. **a**, Representative outpatient platelet count (PLT) trajectories with estimated perRIs using the Gaussian (green) and Bayesian (blue) models, with quintile calibration curves of variance estimates, against a theoretical normal distribution. **b**, Index of individuality (IOI: intra-patient coefficient of variation [CV] / inter-patient CV) for 43 common outpatient laboratory markers. **c**, Mean RMSE, KS statistic, and cross-quintile KS range for the 43 markers, in an evaluation set of 10,000 patients; for the Bayesian, Gaussian models, and popRI as a reference standard. **d**, Number of patients with estimable setpoints (3+ outpatient measurements) for the 43 markers. **e**, Association of the Bayesian model RMSE and KS statistics with IOI. **f**, Bayesian, GMM, and popRI error metrics for PLT stratified by the number of data points used for perRI construction. All error metrics are calculated based on fit against the next isolated outpatient test after model fitting. Theoretical lower bound of RMSE is 1.0. Full descriptions of each test abbreviation are given in **Table 1**. Marker, setpoint, and CV distributions are given in **Supp. Fig. 1-2**.

### Bayesian setpoint estimation is robust to hyperparameter choices

A core challenge of constructing perRIs is that a patient’s physiology can change over time due to natural physiologic shifts. Many common markers exhibit consistent age-related drifts both in pediatric and adult settings^17–19^. Equally, natural events (menopause^9^, etc.), or clinical interventions (long-term medication usage^13^, smoking cessation^7^, etc.) may temporarily or permanently alter homeostatic setpoints. To account for this, we incorporated a temporal weight parameter, *λ*, to allow for gradual setpoint shifts. Through an example patient case, the role of *λ* can clearly be seen, with too high values leading to unstable setpoint estimates, and too low values leading to a loss of adaptability (**Fig. 2a-b**).

**Figure 2.**
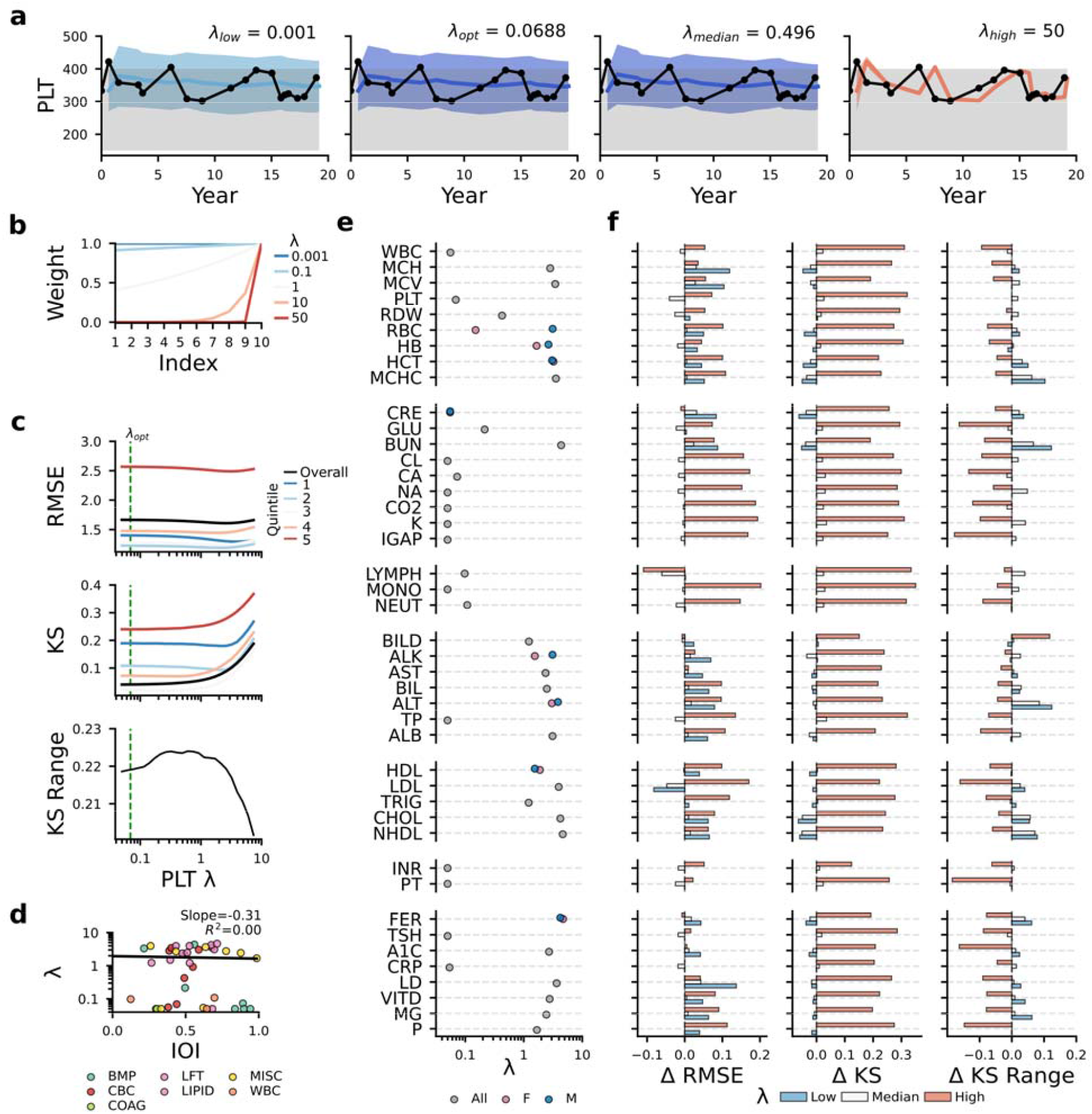
Effect of uneven data weighting on Bayesian model fits. **a**, Representative model fits for a platelet count (PLT) trajectory, under varied weighting of data points, through the memory loss parameter λ. **b**, Examples of resultant data point weighting across 10 measurements, under varying λ. **c**, Error metrics for PLT for a test cohort, and each data quintile, as λ varies. **d**, Association between λ and the marker index of individuality (IOI). **e**, Distribution of optimized λ values across the 43 markers, stratified by sex, and ordered by ascending IOI within each marker group. **f**, Change in error metrics when using the median λ across all 43 markers, and when using extremely high or low λ values, compared to the marker-specific optimum. Error optimization curves for individual markers are given in **Supp. Fig. 3**.

Typically, the choice of *λ* had inverse effects on RMSE and KS, with higher values often allowing for more adaptability to setpoint shifts, at a cost of worsening variance calibration (**Fig. 2c**). Optimization curves for all markers are shown in **Supp. Fig. 3**. The optimized *λ* showed no association with each marker’s individuality (Spearman = -0.05) (**Fig. 2d**). While optimal *λ* values varied across the set of markers, sex-stratified estimates remained closely aligned within each marker (Wilcoxon p = 0.42) (**Fig. 2e**). A cohort-median *λ* achieved performance indistinguishable from marker-specific optima (median paired ΔRMSE =-0.003 [IQR -0.017, 0.010], Wilcoxon p = 0.413) with all markers remaining within 5% of optimal performance (**Fig. 2f**). Notably, the reduction in KS range observed with extremely high *λ* likely reflected uniformly poor calibration across strata rather than improved consistency (**Fig. 2f**).

### Personalized setpoints predict mortality and morbidity before population thresholds are crossed

Setpoints of all 43 laboratory markers were strongly associated with future all-cause mortality (**Fig. 3a, Supp. Fig. 4**) even when limited to within popRI (i.e. normal) values. Higher setpoint variability (as defined by the coefficient of variation [CV]) – indicative of worse homeostatic control – was also a consistent sign of increased all-cause mortality (**Fig. 3a, Supp. Fig. 4**). Bayesian risk signals were systematically stronger than GMM estimates, for setpoint risk (median *Δ*log(HR)_CV_ =0.0175, Wilcoxon p=0.008) and variance-related mortality risk (median paired *Δ*log_CV_ = 0.08, Wilcoxon p=0.004; **Fig. 3a**). Bayesian setpoints and CVs showed increasingly strong risk signatures as more data points were used to estimate each perRI (**Fig. 3b**). Setpoint differences inside the popRI were also associated with substantial risks of future morbidity across widely varied markers and disease states (**Fig. 3c**). Of note, many of these disease states (e.g., iron-deficiency anemia, non-alcoholic fatty liver disease, etc.) reflected over 3-fold risk increases between the highest-risk quartile and remainder of the cohort, similar to the degree of risk stratification seen in many public health screening guideline factors (e.g., family history of disease, age thresholds for cancer screening, etc.)^20,21^.

**Figure 3.**
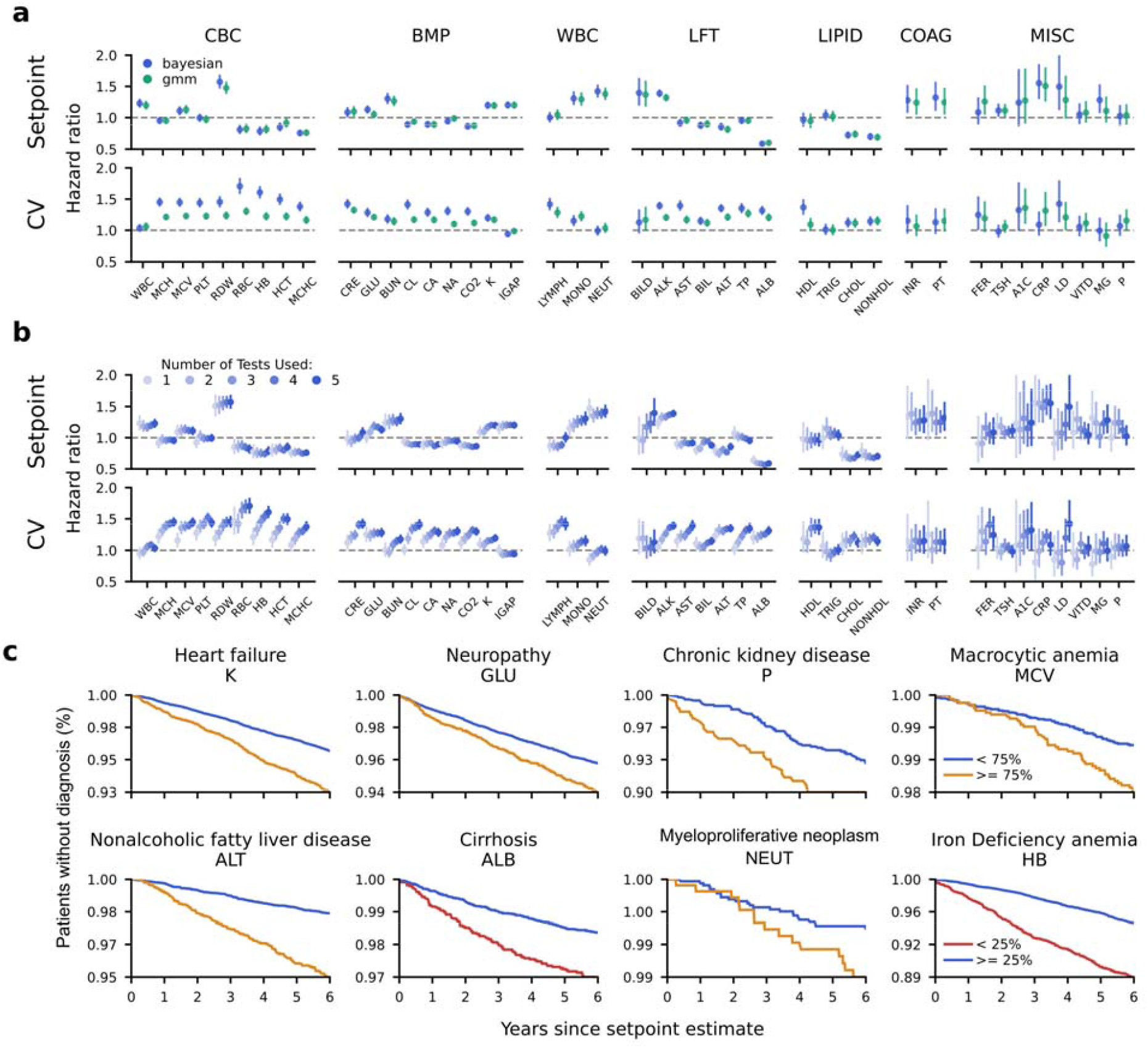
Association of homeostatic setpoints with future morbidity. **a**, Age- and sex-adjusted hazard ratios for setpoint and coefficient of variation (CV) estimates with all-cause mortality, normalized to a one standard deviation increase, using the Bayesian and Gaussian models. **b**, Hazard ratios for setpoint and CV as more data is used to estimate each perRI. **c**, Associations with setpoints and future disease diagnoses (i.e., patients without disease diagnosis), stratified by setpoint quartile. Setpoints in **c**, are estimated using the first five isolated outpatient measurements of each test. All associations in this figure are derived after limiting to setpoints to values within the population reference interval. Results without this constraint are given in **Supp. Fig. 4**.

### Personalized reference intervals complement and enhance existing risk stratification approaches

Clinical laboratory markers encode two axes of patient risk simultaneously: the risk induced by the absolute physiologic value (e.g., elevated hematocrit leads to higher blood viscosity, increasing cardiovascular disease risk), and the risk induced by deviation (e.g., a decrease in hematocrit may signal increased red cell destruction). To capture these two risks, we stratified outpatient cohorts based on whether their presenting marker was outside the popRI (absolute risk) and/or outside the perRI (deviation risk) across multiple major disease cohorts with well-established clinical marker associations (acute kidney injury [AKI] and creatinine [CRE]; leukemia and white blood cell count [WBC]; and hypothyroidism and thyroid stimulating hormone [TSH]) (**Fig. 4a-d**). In these cohorts, disease development was often preceded by gradual decline, meaning patients deviated from their perRI years before clinical diagnosis (which often coincided with the popRI deviation) (**Fig. 4a**). Given this, deviation from the popRI and perRI often denoted similar magnitudes of risk (**Fig. 4b**). However, these risk signals are highly complementary, as patients who deviated from both ranges simultaneously showed far greater risk elevations (**Fig. 4c**) than patients who deviated from either interval alone. Compared to patients within both intervals, patients outside both intervals exhibited 7-fold increased 2-year diagnosis risk for AKI (0.3% vs 2.8%, p<0.001), 8-fold for leukemia (0.1% vs 0.8%, p<0.001), and 3-fold for hypothyroidism (2.8% vs 8.3%, p<0.001) (**Fig. 4d**). Strong risk stratification was seen for deviations inside the popRI but outside the perRI even in cases such as hypothyroidism, where TSH outside the popRI is often the primary trigger for additional diagnostic work-up, such as free thyroxine testing (**Supp. Fig. 5c**)

**Figure 4.**
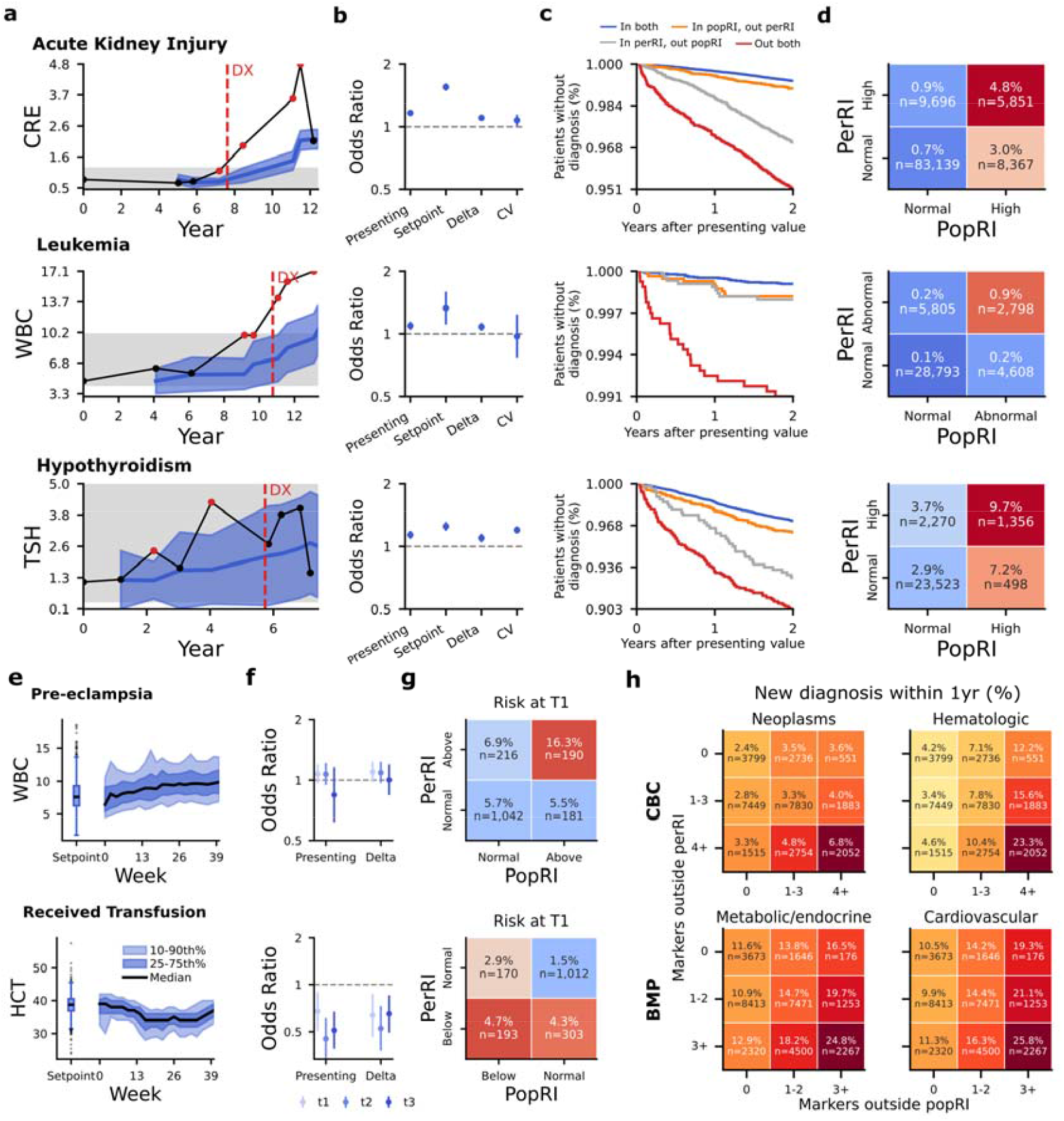
Prognostic value of personalized reference intervals. **a**, Examples of perRI construction and outpatient monitoring for three cohorts with well-established marker-disease associations (creatinine and acute kidney injury; white cell count and leukemia; and thyroid stimulating hormone and hypothyroidism). **b**, Odds ratios of the presenting outpatient value, setpoint and CV, and delta (difference between setpoint and presenting value) for attaining each diagnosis within the next two years. **c-d**, Kaplan-Meier curves (**c**), and two-year diagnosis rates (**d**) for patients, stratified by whether their presenting value is inside the population and/or personalized reference interval. **e**, Distribution of pre-pregnancy setpoints and marker values across gestation in a large pregnancy cohort. **f**, Odds ratios for development of pre-eclampsia or for needing a transfusion during delivery, for the presenting value or delta (change from setpoint) across trimesters. **g**, Risk of negative outcome stratified by whether the presenting value in trimester 1 is within the population and/or personalized reference interval. **h**, Risk of new diagnosis within the following year, stratified by how many of a panels’ markers are outside the population and/or personalized reference interval. Results for hypothyroidism use a composite outcome based on ICD code diagnosis, or presence of free thyroxine levels below 0.6. Results for each of these composites separately are given in **Supp. Fig. 5**. Results in **e-g** limited to each patient’s first pregnancy are given in **Supp. Fig. 6**.

This joint value of popRIs and perRIs generalized to dynamic physiologic settings, such as during pregnancy (**Fig. 4e-g**). During trimester 1, joint deviation from the popRI and perRI conveyed a 3-fold increased risk of pre-eclampsia (WBC, 16.3% vs 5.7%, p < 0.001) and of future transfusion need (HCT, 4.7% vs 1.5%, p = 0.008) compared to patients inside both intervals (**Fig. 4f-g**). Risk stratification remained strong across future trimesters, with the deviation from setpoint providing similar but complementary risk signals to the presenting marker **(Fig. 4f**). Similar directional effects were observed when analyses were restricted to first pregnancies only^22^ (**Supp. Fig. 6**).

The risks of perRI deviations strongly aggregated over multiple marker panels (**Fig. 4h**). Outpatients who presented with multiple blood count or metabolic panel tests outside the popRI and perRIs were at extremely elevated one-year risk of neoplasms (CBC, 6.8% vs 2.4%, p<0.001), hematologic disorders (CBC, 23.3% vs 4.2%, p<0.001), metabolic/endocrine disorders (basic metabolic panel [BMP], 24.8% vs 11.6%, p<0.001), and cardiovascular diseases (BMP, 25.8% vs 10.5%, p<0.001). This is an important finding given that most routine clinical markers are measured as part of multi-marker test panels, meaning joint test interpretation can be readily utilized across most clinical contexts.

### Personalized reference intervals can help predict treatment response

Given the strong associations of perRIs with future diagnoses, we investigated whether they could aid in clinical treatment decisions. As a targeted use case, we considered all patients who were treated with an iron sucrose infusion. We constructed their hemoglobin (HB) perRI using all data up to one year pre-infusion and considered their pre-treatment HB (0-60 days pre-infusion) and post-treatment HB (2-6 months post-infusion) (n: 492) (**Fig. 5a-b**). Benchmarking against the HB setpoint revealed clear trends, with patients exhibiting HB declines in the year prior to iron infusion, and on average a gradual HB increase following infusion, reaching their pre-disease setpoint between 3-4 months post treatment (**Fig. 5c**). The HB drop from setpoint to pre-treatment was more predictive of the post-treatment HB increase (“response”) than the pre-treatment value (R^2^ = 0.29 vs 0.23, p = 0.001) (**Fig. 5d**). The HB drop was positively associated with the magnitude of treatment response regardless of the patient’s pre-treatment HB (**Fig. 5e**). Considered jointly, patients who were outside both the popRI and perRI were twice as likely to exhibit a >1 g/dL HB treatment response than patients inside both intervals (35.8% vs 76.2% for females, p < 0.001; 65.3% vs 25.0% for males, p=0.01). Considered separately, patients who deviated from the perRI showed substantially higher rates of >1 g/dL HB response than patients who deviated from the popRI (76.8% vs. 42.1% for females, p < 0.001; 64.0% vs 41.7% for males, p=0.030) (**Fig. 5f**). This suggests that perRIs are substantially more predictive of treatment responses than currently used popRIs.

**Figure 5.**
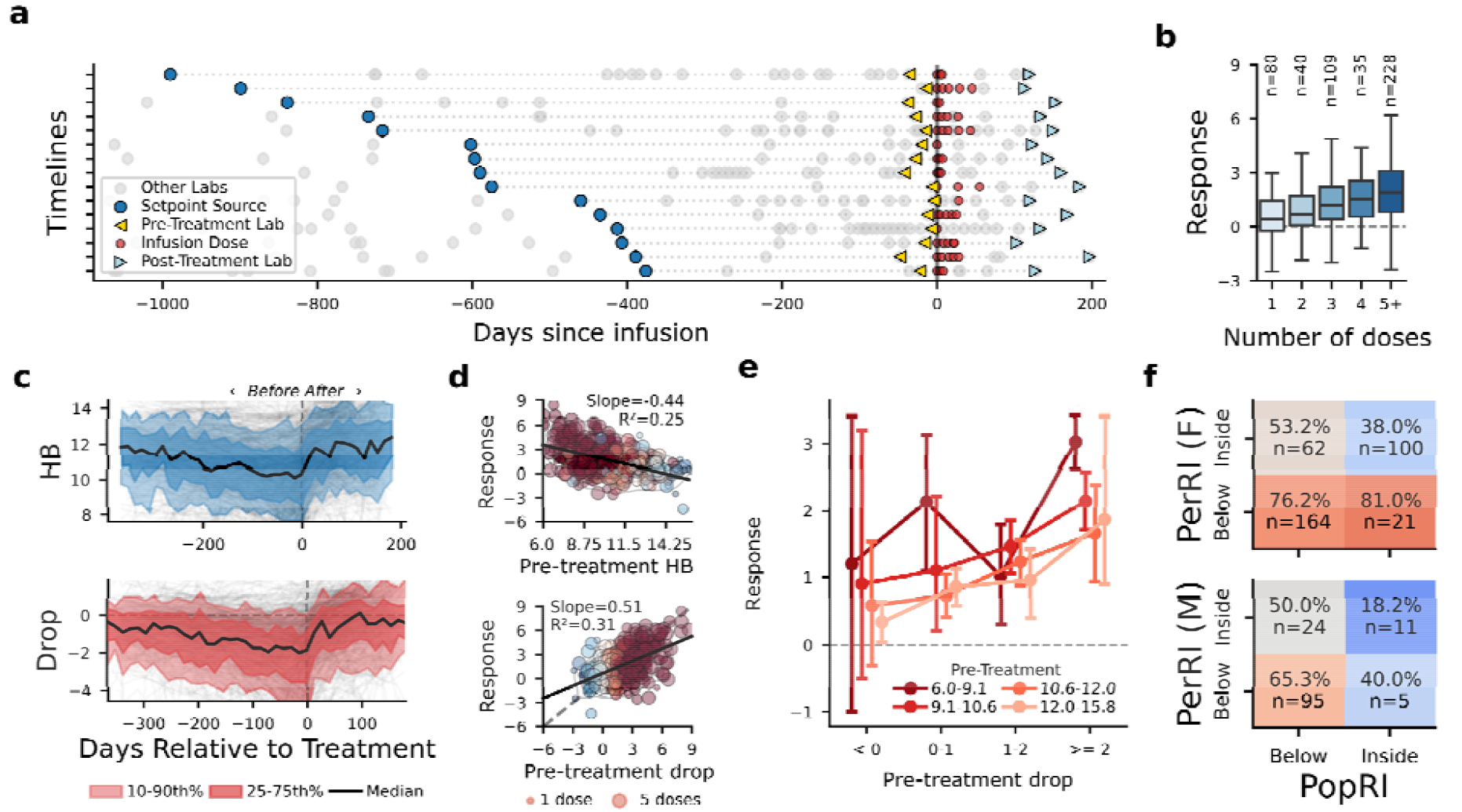
Value of personalized reference intervals for iron infusion prognostication. **a**, Time points of hemoglobin (HB) labs, setpoint source, iron infusion codes, and pre- and post-treatment HB measurement in an iron infusion cohort. **b**, Distribution of the post-treatment HB increase (“response”) across over the number of doses received across the cohort. **c**, Response trajectories for HB, and the HB drop from setpoint, pre- and post-treatment. **d**, Association of pre-treatment HB and HB drop with the response. **e**, Association between HB drop and response across presenting HB values. **f**, Likelihood of a >1g/dL HB response by whether the pre-treatment HB was within the population or personalized reference interval.

## Discussion

Clinical laboratory markers reflect individually regulated traits but are interpreted with one-size-fits all population reference intervals. We showed that Bayesian inference can be used to construct accurate and clinically relevant personalized reference intervals, using each patient’s pre-existing health record. This shifts laboratory test interpretation from identifying abnormal values within a population to detecting deviations from an individual’s personalized benchmarks.

The highly heritable nature of most laboratory markers has been well-established over short time periods through efforts such as from the European Federation of Laboratory Medicine^23^. In recent work, we showed that this regulation persisted over decades for the CBC^16^. Our results extend these findings to a far broader suite of laboratory tests, reflecting markers of immune, metabolic, cardiovascular, and renal function. Of the 43 most common laboratory test markers by volume at our center, 33 had IOIs below 0.8, suggesting benefit from perRI usage. Clinicians have long recognized that some laboratory values must be interpreted considering a patient’s history, but this process often depends on informal trend review and clinical experience instead of robust quantitative processes^24^. Given this, our framework has the potential to genuinely revolutionize routine laboratory medicine, shifting a central paradigm of how nearly all routine clinical tests are interpreted.

Importantly, we showed that patient-specific benchmarks can be constructed from extremely limited data. Health record availability will inevitably vary across populations due to differences in patients’ healthcare exposures, testing histories, and due to limited data sharing across separate healthcare centers^25^. As such, any viable approach to personalized test interpretation must be data-adaptive and effective with only minimal prior testing. Our Bayesian approach solves this problem elegantly, reverting to the popRI in the presence of no prior data, and otherwise providing a gradually refining perRI estimate. Our findings also suggest that clinical value can be derived even from coarse perRI estimates that use only 2-3 prior patient tests. This means the benefits of our approach are not limited to patients with the most extensive medical histories but can instead be broadly beneficial across the population and across ostensibly healthy patient states^26^.

A related benefit of perRIs may lie in enhancing longitudinal patient monitoring. For markers with significant age effects, popRIs are often age-stratified, with different intervals across pediatric and elderly ages. While it is important to reflect population-level physiologic shifts, this strategy can inadvertently normalize physiological decline, reducing sensitivity for disease detection^3^. Similarly, age-stratified popRIs are typically implemented as step-functions, with sudden jumps at specific ages (i.e. alkaline phosphatase often has separate popRIs for 18-24, 25-34, 35-44 years old, etc., see **Supp. Table 1**). This may reduce diagnostic sensitivity as it does not reflect the continuous nature of physiologic drift and context^27^. Anchoring interpretation to an adaptive personal baseline instead inherently captures these physiological shifts over time while still reflecting the shifting definition of health as populations age. What follows is more accurate and sensitive detection of dysregulations that exceed the bounds of normal age-related drift and biologic fluctuation.

Across nearly all routine markers, setpoints and CVs were strongly associated with mortality and morbidity. Of note, elevated variability was consistently associated with increased mortality, even when CVs were estimated from as few as three measurements, suggesting CV estimates capture true patient-specific signals about homeostatic control^28^. Similar to our prior report for the blood count, non-hematologic marker setpoints retained strong mortality associations when limited to values inside the popRI^16^. This reinforces the concept that marker-risk associations are continuous, and that popRIs do not inherently reflect a significant risk cliff, particularly for general patient outcomes.

The key finding of our study is that popRIs and perRIs capture broadly orthogonal aspects of patient risk. Compared to popRIs, perRIs can more sensitively distinguish biologic fluctuations from pathophysiologic declines, allowing for early detection of emergent disease. Conversely though, perRIs cannot detect risks due to longitudinally stable dysfunction or abnormality – whether driven by genetics, lifestyle factors, or comorbid disease. Therefore, popRIs and perRIs are best viewed as complementary approaches to patient prognostication. Evidenced across widely varied markers and disease settings, we consistently find that the highest risk patients are those who deviate from both population and personal references where risk stratifications exhibited across our results are often greater than 3-fold across the four major groups (inside/outside popRI or perRI). This magnitude is similar to commonly implemented disease screening factors, such as family history^21^. This suggests that a joint popRI and perRI approach to laboratory test interpretation could significantly aid outpatient disease screening efforts.

### Limitations

While intuitive and robust across markers, our results are limited by the use of retrospective data from a single major medical center. As such, prospective external validation is needed. Similarly, while results show strong potential for clinical value, use of perRIs to inform clinical care must first be validated in appropriate clinical settings (randomized trials, etc.), a clear avenue for future research. We also recognize that for widespread usage of perRIs, they must be carefully integrated into existing EHR infrastructure, which is another major avenue for future work.

### Conclusions

Collectively, our findings show how Bayesian inference can allow for construction of personalized interpretations of routine laboratory tests. Combined use of personalized and population reference intervals can greatly enhance diagnostic and prognostic sensitivity across a wide variety of tasks. This has the potential to transform how laboratory test interpretation is approached, driving forward precision medicine that is grounded in routinely collected data.

## Methods

### Data Source and Cohort Construction

We conducted a retrospective analysis of longitudinal laboratory data derived from the University of Washington Medicine (UWM) EHR. We considered all adult patients with at least three isolated outpatient tests for one or more of 43 markers selected based on the frequency of their use in outpatient medicine (defined in **Table 1**). Measurements were considered isolated if they were separated by more than 90 days from both the preceding and subsequent measurement of the same marker, reducing the influence of acute illness, short-term monitoring, and transient physiologic perturbations. For all patients, we collated demographics, laboratory test orders and results, International Classification of Diseases (ICD) coded diagnoses, and medication administration records. Laboratory test data was taken from as far back as UWM’s medical record digitization extends (the earliest isolated laboratory marker was from 2008-06-16), up until the data collection date (2025-06-17). We considered all markers from the complete blood count (CBC), white blood cell differential (WCD), basic metabolic panel (BMP), liver function tests (LFT), lipid panel (LIPID), and coagulation studies (COAG), as well as selected set of quantitative, high testing volume markers of inflammation, tissue injury, thyroid function, glycemic status, iron stores, vitamin D status, and mineral status (MISC). A full list of markers and their popRIs are given in **Table 1**.

### Longitudinal Modeling Framework

We used a Bayesian inference framework to estimate setpoints. We assumed that isolated outpatient measurements for a patient and marker were Gaussian-distributed around an individual, time-varying setpoint *μ*_*t*_, with variance 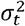. Markers with skewed population-level distributions were log-normalized prior to setpoint estimation (denoted in **Table 1**). The model was initialized with weakly informative uniform priors, *P*(***θ***_*t*_), over physiologically plausible values. Specifically, *μ*_*t*_ was assigned a uniform prior over an extended population range, set as the popRI midpoint ± 2 × *POPRI*_*Width*_ (e.g., if the WBC popRI is 4-10, the range would be 7 ± 6: 1-13). *α*_*t*_ was assumed to be uniformly distributed between 0 and the marker’s inter-patient standard deviation, estimated from the width of the population reference interval. From this prior, estimates of *μ*_*t*_ and *α*_*t*_ were iteratively refined using Bayes theorem:

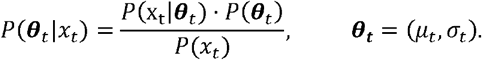

For the likelihood, we used a Gaussian distribution. We implemented recency weighting by raising each measurement’s Gaussian likelihood contribution to an exponential decay weight through a marker-specific parameter *λ*:

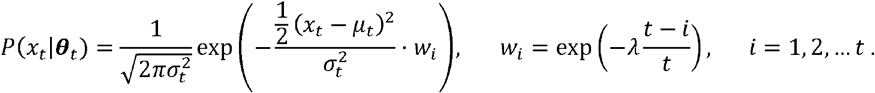

This model was optimized using direct calculation of the likelihood integrals over a 201×201 joint distribution grid of *μ*_*t*_ and *α*_*t*_ values. For all patients, models were fit using the first *t* measurements and evaluated based on their fit against the *t+1*^th^ value. Error was calculated based on the root mean squared error (RMSE) between *μ*_*t*_ and x_t+1_, and to enable comparison of markers with different natural ranges, RMSE was scaled by the median intra-patient standard deviation, where the theoretical lower bound on RMSE is 1.0. Calibration was assessed by computing the Kolmogorov-Smirnov (KS) statistic, to measure the maximum deviation of patient Z-scores (values normalized by their perRI setpoint and standard deviation) from a theoretical standard normal distribution.

To account for overestimates in some patients being cancelled out by underestimates in other patients (artificially reducing the KS statistic), we also considered the maximum difference in KS values across each quintile of x_t+1_ values, with this being denoted the KS range. Model performance was compared to that achieved by our previously described Gaussian mixture model^16^, and to the error achieved by the population reference range (i.e., *μ*_*t*_ = popRI center, *α*_*t*_ is derived from the popRI, assuming it’s a 95% confidence interval).

To optimize *λ*, we considered a set of 10,000 randomly chosen patients for each marker. We optimized *λ* via differential evolution (population size=8, tolerance=0.001) and 5-fold cross-validation. Cross-validation folds were partitioned at the patient level. Optimization minimized a composite objective consisting of normalized RMSE, the KS statistic, and KS range. To select a single hyperparameter configuration for deployment, the independently optimized candidate from each cross-validation fold was evaluated across all validation folds, and the candidate with the lowest mean validation loss was retained. *λ* was optimized over a bounded continuous interval on the log scale (log (*λ*) ∈ [−3,4] corresponding to *λ* ∈ [0.05,54.6]

### Setpoint outcome associations

We evaluated associations between individualized setpoints and clinical outcomes using time-to-event analyses. For each patient and laboratory marker, setpoints were anchored at a fixed longitudinal index to obtain a single estimate of setpoint mean (succinctly referred to as the setpoint) and coefficient of variation (*CV* = *σ*/ *μ*). Patients with a diagnosis of the outcome of interest prior to the anchoring date were excluded. Associations with all-cause mortality were evaluated using Cox proportional hazards models adjusted for age and sex. To account for marker scale, hazard ratios were reported per one standard deviation increase. To evaluate the temporal stability of inferred associations, analyses were replicated using setpoint estimates based on different numbers of prior test values.

Associations with future disease diagnoses were evaluated using ICD-9 and ICD-10 codes. Patients were included if they had a fifth setpoint estimate observed between 2014 and 2019, which served as the anchoring date for follow-up. Diagnosis-specific Kaplan–Meier analyses were performed by stratifying patients into marker-specific risk groups based on setpoint distributions, with directionality selected to reflect expected disease physiology. Patients were censored at first diagnosis, death, or six years after the anchoring date, whichever occurred first.

The following ICD codes were used for each diagnosis in the Kaplan–Meier analyses: iron deficiency anemia (ICD-9: 280; ICD-10: D50); cirrhosis (ICD-9: 571.2, 571.5, 571.6; ICD-10: K74.3, K74.4, K74.5, K74.6); nonalcoholic fatty liver disease (ICD-9: 571.8; ICD-10: K76.0, K75.81); rheumatoid arthritis (ICD-9: 714; ICD-10: M05, M06); chronic kidney disease (ICD-9: 585; ICD-10: N18); neuropathy (ICD-9: 356.9; ICD-10: G62.9); venous thromboembolism (ICD-9: 415.1, 451, 453; ICD-10: I26, I80, I82); heart failure (ICD-9: 428; ICD-10: I50). Each ICD code includes all sub-codes. Hypothyroidism diagnosis was additionally considered as any presenting free thyroxine (T4FR) test result less than 0.6.

### Personalized Reference Interval Analyses

To assess the clinical utility of perRI during individual clinical encounters, we examined three conditions with distinct temporal dynamics over a two-year window: acute kidney injury (AKI) and creatinine (CRE); leukemia and white cell count; and hypothyroidism and thyroid stimulating hormone [TSH]. Patients were identified at their third setpoint estimate and required to remain free of the outcome diagnosis for at least one year. The subsequent outpatient laboratory value after this one-year washout period was then treated as the presenting measurement.

We evaluated whether setpoint-derived features provided prognostic information beyond the absolute presenting value using univariate logistic regression within the two-year prediction window. For each predictor (presenting value, delta=presenting-setpoint, setpoint, and setpoint CV), a separate model was fit, adjusted for age and sex. Patients were additionally classified into four mutually exclusive groups based on whether their presenting values where in the popRI, or perRI (or both or neither). Kaplan–Meier analyses were used to evaluate time-to-diagnosis differences across groups, censored at two years after the presenting test date or date of death, whichever occurred first.

To assess joint interpretation in a case of physiological adaptation, we performed a complementary analysis in a pregnancy cohort stratified by trimester: T1 (0–13 weeks), T2 (14–27 weeks), and T3 (28 weeks through delivery). Individualized setpoints were estimated using at least three historical pre-pregnancy test measurements. For each trimester, the presenting laboratory value was defined as the measurement closest to the midpoint of the trimester interval. Pre-eclampsia was identified using ICD-10 code O14, and transfusion was based on recorded infusion of blood products during pregnancy or delivery.

### Multi-Marker Burden Heatmaps

To assess whether personalized and population reference intervals capture complementary risk dimensions across panels, we identified patients at their fifth individualized setpoint estimate and extracted their next outpatient CBC and BMP panel values after a one-year washout period. For each marker, patients were classified as inside or outside the popRI and inside or outside their perRI. Patients were then binned by the count of markers outside the popRI (0, 1–3, or 4+ for CBC; 0, 1–2, or 3+ for BMP) and the count outside the perRI. Within each cell, we computed the proportion of patients acquiring a new diagnosis across four clinically relevant outcome categories: solid and hematologic neoplasms (ICD10: C00-C96), hematologic disorders (D50-D89), metabolic and endocrine diseases (E00-E89), and cardiovascular diseases (I00-I99). New diagnoses were defined as any new whole number ICD10 code (i.e., C01, C02, etc.) excluding any codes present in the patient’s record prior to the presenting measurement.

### Iron Infusion Analysis

To evaluate whether deviation from individualized setpoints contextualized treatment response, we analyzed hemoglobin responses in all UWM patients receiving an intravenous infusion of iron sucrose. Infusions were identified from medication administration records and grouped into treatment courses (any set of infusions with no more than 60 days separation). To minimize confounding, we considered only each patient’s first recorded iron infusion treatment course. Hemoglobin perRIs were estimated using all adult isolated HB tests from more than one year prior to infusion. Patients with fewer than three valid HB measurements in this range were excluded. Pre-treatment hemoglobin (HB_pre_) was defined as the laboratory value closest to treatment initiation within the preceding 60 days. Post-treatment hemoglobin (HB_post_) was defined as the earliest measurement 60–180 days after course completion and prior to any subsequent treatment course. To evaluate the value of setpoints for iron infusion prognostication, we quantified drop (setpoint −HB_pre_) and response (HB_post_ − HB_pre_). Logistic regression was used to evaluate whether deviation from setpoint improved prediction of large hemoglobin response (>1 g/dL) beyond baseline hemoglobin. Nested models comparing HB_pre_ alone versus HB_pre_ and hemoglobin drop were evaluated using likelihood-ratio tests for incremental model fit.

### Statistical Analysis

All statistical analyses were performed independently for each laboratory marker unless otherwise specified. Paired comparisons of performance metrics between models were evaluated using the Wilcoxon signed-rank test across markers. Associations between marker-level quantities were assessed using Spearman rank correlation. Survival analysis was performed using Kaplan–Meier curves, with statistical significance calculated using the log-rank test. Time-to-event associations were modeled using Cox proportional hazards regression, with hazard ratios reported per one standard deviation increase in the relevant setpoint parameter. Differences in event rates between groups were tested using the chi-squared test or Fisher’s exact test. All statistical tests were two-sided, with significance defined as Holm-adjusted p < 0.05 unless otherwise specified. All analyses were performed using Python with scipy, statsmodels, lifelines, and numpy.

## Acknowledgements

We thank the UW Department of Laboratory Medicine & Pathology’s Informatics team for assistance in data acquisition.

## Ethics

This study was performed under a waiver of informed consent due to minimal risk, with a protocol approved by the University of Washington Medicine Institutional Review Board.

## Code Availability

Code for the construction of setpoint and personalized reference interval estimates using a Bayesian inference model are available at https://github.com/cnmnzhang/perRI.

## Data Availability

Due to limitations on sharing of protected health information, individual patient data cannot be shared.

## Author contributions

BHF and CZ conceived of the study. CZ performed primary data analysis, with contributions from YC, AJ, and EL. SS and BDL provided clinical input. All authors contributed to data interpretation, and manuscript writing.

