## Supplemental File 1 for "Personalized clinical reference intervals for routine precision medical care"

**SUPPLEMENTARY FILE 1**

**Personalized clinical reference intervals for routine precision medical care**

Cindy Zhang^1,2^, Ya-Lin Chen^1,2^, Attila Jamilov^1^, Elvin Liu^1^, Raj Shree^3^, Barbara Lam^4,5^, Brody H Foy^1,6,7,^*

^1^Department of Laboratory Medicine & Pathology, University of Washington Medicine, Seattle, USA.

^2^Department of Biomedical Informatics & Medical Education, University of Washington Medicine, Seattle, USA.
^3^Division of Maternal-Fetal Medicine, Department of Obstetrics & Gynecology, University of Washington Medicine, Seattle, USA
^4^Division of Hematology & Oncology, Department of Medicine, University of Washington Medicine, Seattle, USA
^5^Division of Hematology & Oncology, Fred Hutchinson Cancer Center, Seattle, USA
^6^Department of Bioengineering, University of Washington, Seattle, USA.
7Brotman Baty Institute for Precision Medicine, Seattle, USA.

Contents

- Supplementary Table 1
- Supplementary Figures 1-6

Supplementary Tables

**Supplementary Table 1 | Age-dependent population reference intervals for ALK at UWM.**

| **Population reference interval** | | |
| --- | --- | --- |
| **Age** | **Male** | **Female** |
| 0-9y | 111-281 | 115-324 |
| 10-11y | 132-366 | 115-324 |
| 12-13y | 89-284 | 119-426 |
| 14-17y | 43-226 | 72-400 |
| 18-24y | 26-98 | 42-136 |
| 25-34y | 25-100 | 35-109 |
| 35-44y | 25-112 | 36-122 |
| 45-54y | 32-121 | 39-139 |
| 55-64y | 31-132 | 37-159 |
| 65-74y | 38-172 | 36-161 |
| 75y + | 49-199 | 52-227 |

Supplementary Figures

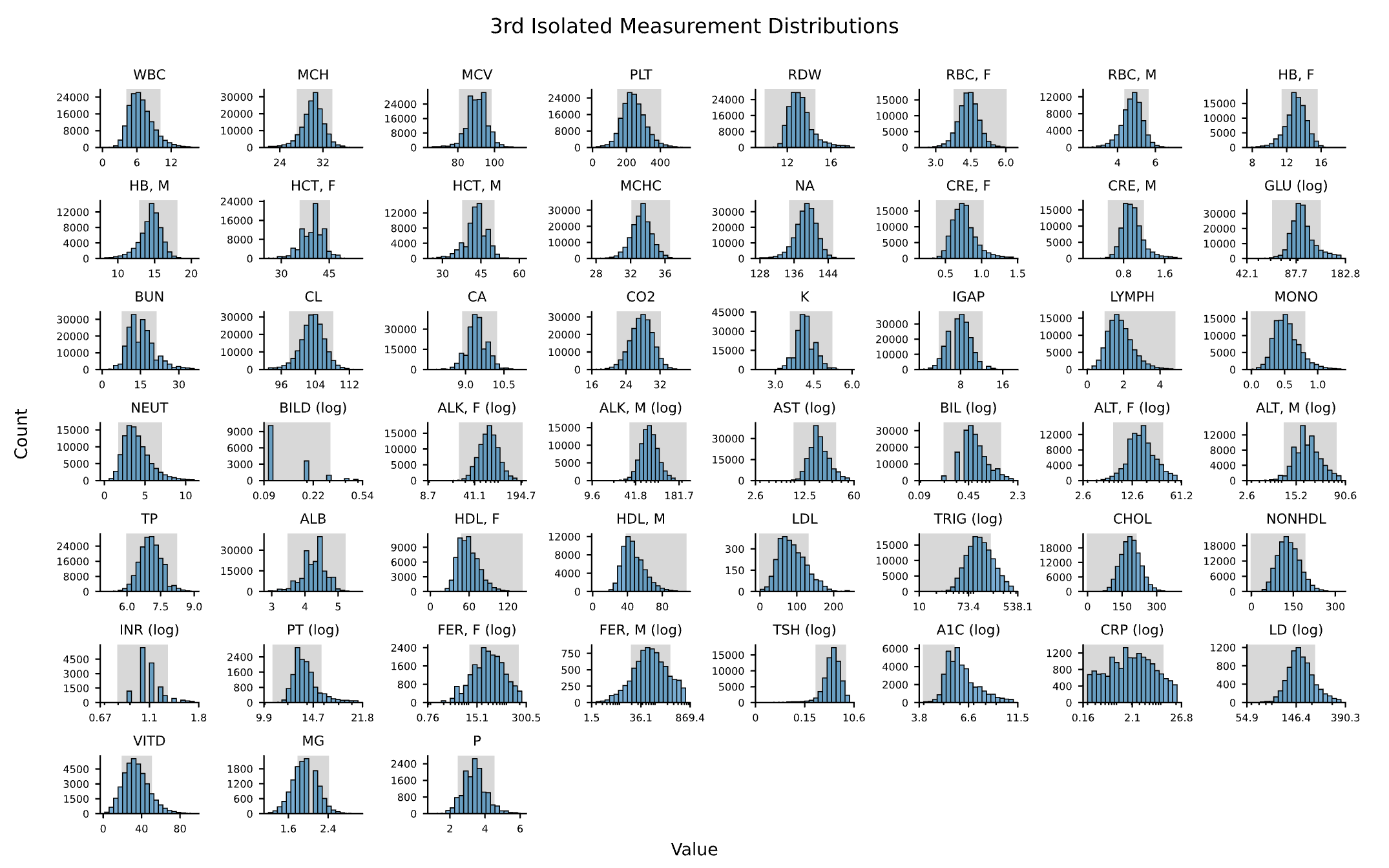

**Supplementary Figure 1 | Marker-level 3^rd^ isolated measurement distributions across the full cohort.** Markers optimized on the log scale are displayed with a log scaled x-axis, and sex-stratified markers are shown separately for female and male patients.

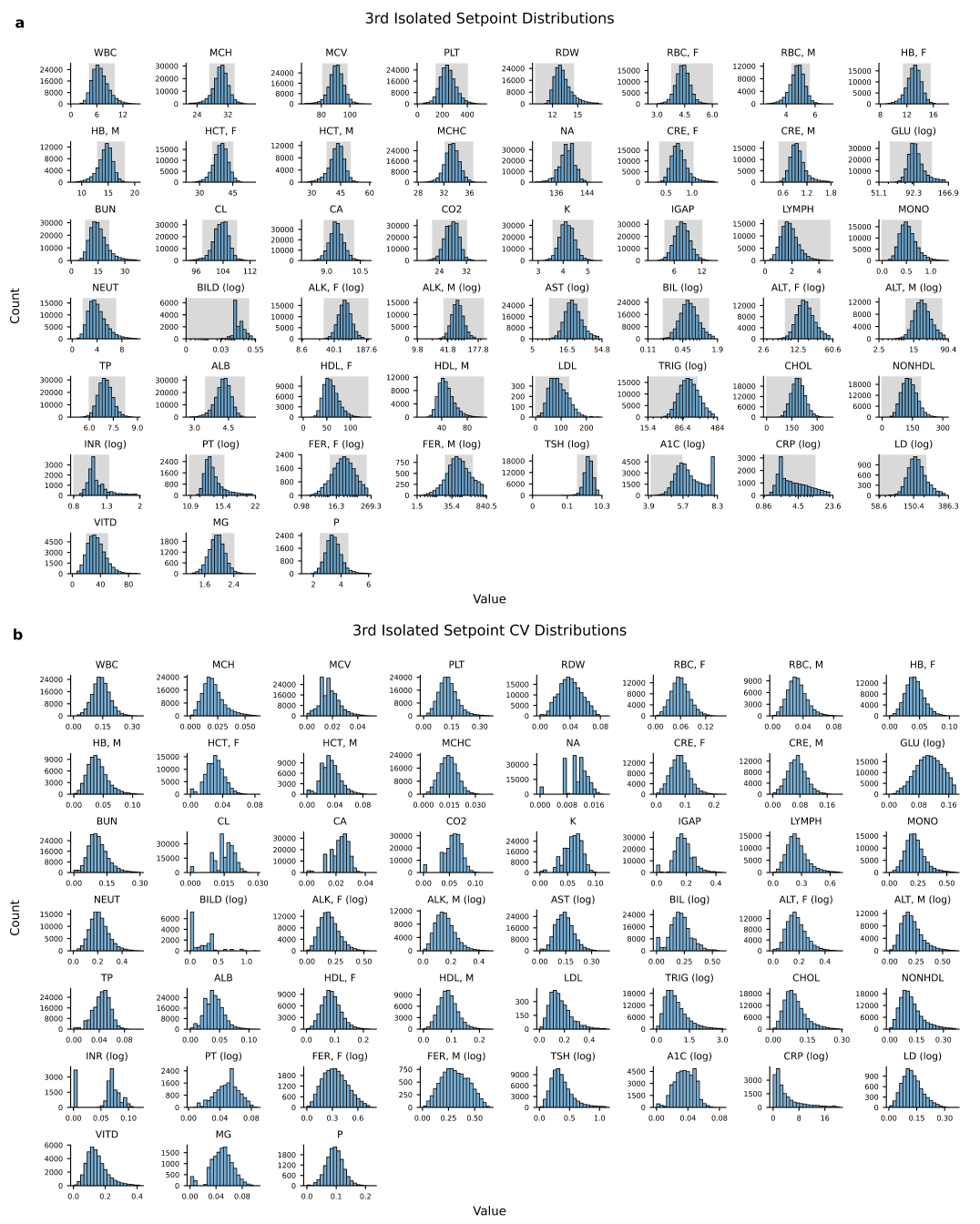

**Supplementary Figure 2. Distribution of setpoint means and coefficients of variation.** Panels use the same marker ordering and sex stratification as **Supp. Fig. 1.** All values were generated using the first three isolated outpatient test results for each patient and marker.

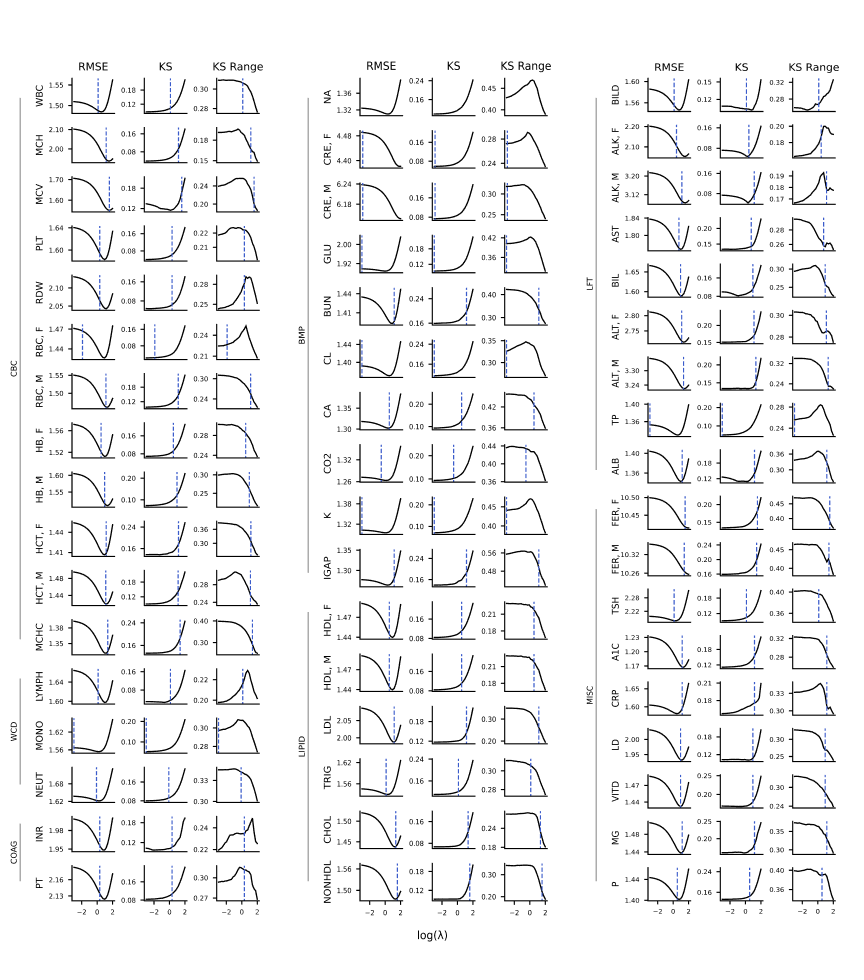
**Supplementary Figure 3.** Error curves across varying lambda values, shown with x-axis on log scale, for the 43 laboratory markers. RMSE is scaled by median intra-patient standard deviation. Dashed vertical reference lines denote grid-based optimal values.

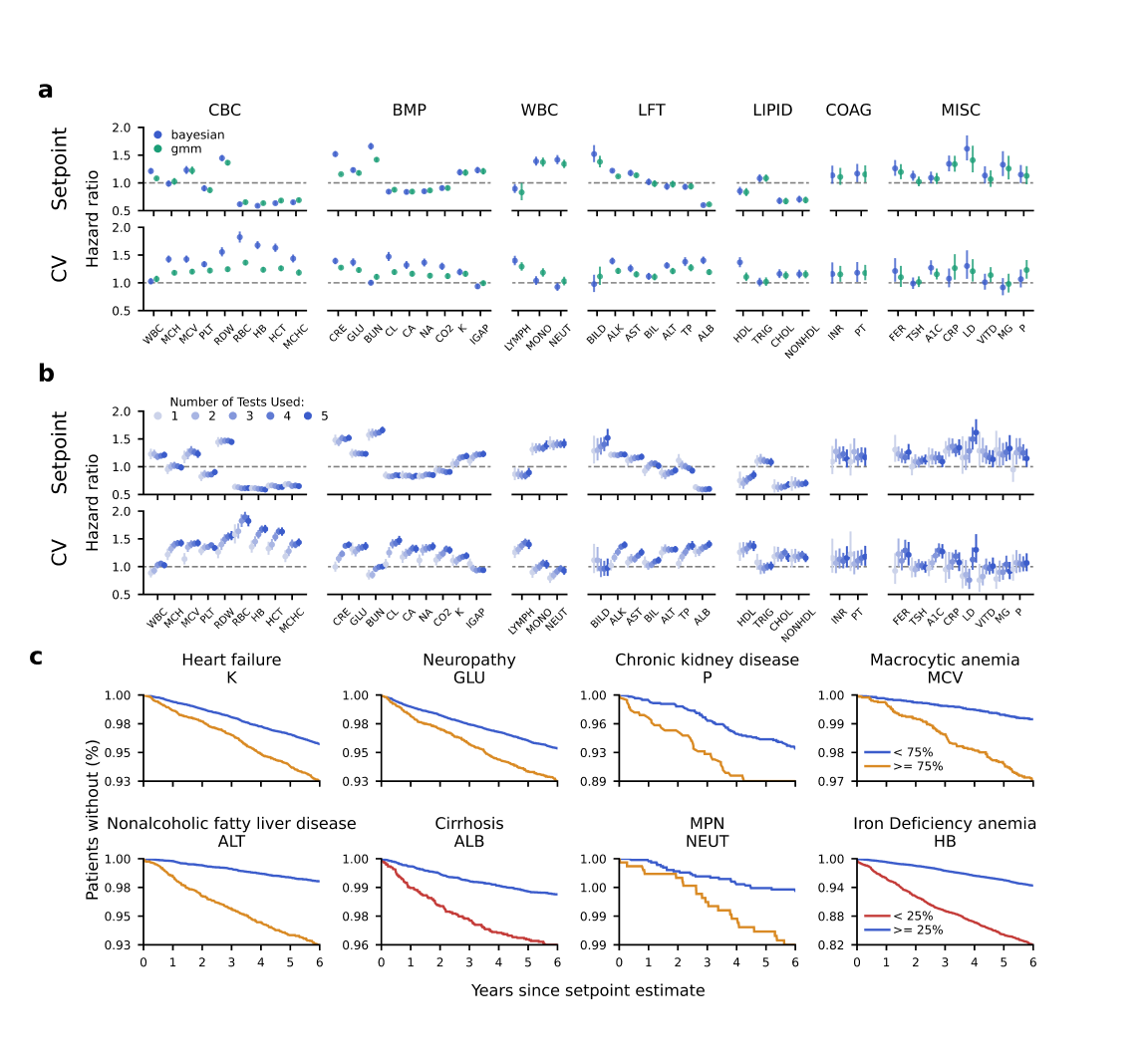
**Supplementary Figure 4 | Setpoint associations with mortality and/or morbidity without restricting to values inside population reference interval.** MPN: Myeloproliferative neoplasm. All lab tests abbreviations are defined in **Table 1**.

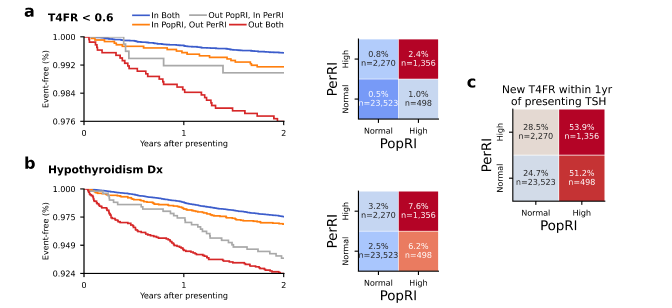
**Supplementary Figure 5 | Consistency of TSH personalized-risk stratification across hypothyroidism outcome definitions.** **a-b,** Kaplan-Meier curves and two-year outcome rates for patients, stratified by whether their presenting value is inside the population and/or personalized reference interval. **(a)** defines outcome of exceeding lower population reference intervals (T4FR < 0.6) and **(b)** Incident hypothyroidism diagnosis codes. **c,** Downstream T4FR ascertainment heatmaps compare subsequent first-time T4FR test order rates across PopRI and PerRI TSH strata for the TSH cohort.

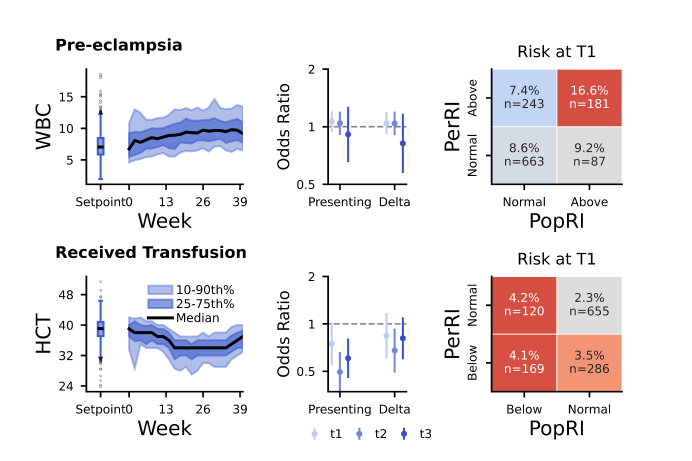

**Supplementary Figure 6 | Prognostic value of perRIs in the first pregnancy.** Results are a replication of results from **Fig. 4**, after limiting to each patient’s first pregnancy.
